# Human norovirus (HuNoV) GII RNA in wastewater solids at 145 United States wastewater treatment plants: Comparison to positivity rates of clinical specimens and modeled estimates of HuNoV GII shedders

**DOI:** 10.1101/2023.05.02.23289421

**Authors:** Alexandria B. Boehm, Marlene K. Wolfe, Bradley White, Bridgette Hughes, Dorothea Duong, Niaz Banaei, Amanda Bidwell

## Abstract

**Background:** Human norovirus (HuNoV) is a leading cause of disease globally, yet actual incidence is unknown. HuNoV infections are not reportable in the United States, and surveillance is limited to tracking severe illnesses or outbreaks. Wastewater monitoring for HuNoV has been done previously and results indicate it is present in wastewater influent and concentrations are associated with HuNoV infections in the communities contributing to wastewater. However, work has mostly been limited to monthly samples of liquid wastewater at one or a few wastewater treatment plants (WWTPs).

**Objective:** The objectives of this study are to investigate whether HuNoV GII preferentially adsorbs to wastewater solids, investigate concentrations of HuNoV GII in wastewater solids in wastewater treatment plants across the county, and explore how those relate to clinical measures of disease occurrence. In addition, we aim to develop and apply a mass-balance model that predicts the fraction of individuals shedding HuNoV in their stool based on measured concentrations in wastewater solids.

**Methods:** We measured HuNoV GII RNA in matched wastewater solids and liquid influent in 7 samples from a WWTP. We also applied the HuNoV GII assay to measure viral RNA in over 6000 wastewater solids samples from 145 WWTPs from across the United States daily to three times per week for up to five months. Measurements were made using digital droplet RT-PCR.

**Results:** HuNoV GII RNA preferentially adsorbs to wastewater solids where it is present at 1000 times the concentration in influent. Concentrations of HuNoV GII RNA correlate positively with clinical HuNoV positivity rates. Model output of the fraction of individuals shedding HuNoV is variable and uncertain, but consistent with indirect estimates of symptomatic HuNoV infections in the United States.

**Significance:** These findings support the utility of wastewater solids as a matrix for community infectious disease surveillance.

**Impact Statement:** Illness caused by HuNoV is not reportable in the United States so there is limited data on disease occurrence. Wastewater monitoring can provide information about the community spread of HuNoV. Data from wastewater can be available within 24 hours of sample receipt at a laboratory. Wastewater is agnostic to whether individuals seek medical care, are symptomatic, and the severity of illness. Knowledge gleaned from wastewater may be used by public health professionals to make recommendations on hand washing, surface disinfection, or other behaviors to reduce transmission of HuNoV, or medical doctors to inform clinical decision making.

## Introduction

Human norovirus (HuNoV) is a leading cause of gastrointestinal illness globally^1^, yet its actual disease burden is largely unknown. This is not only because some infections are asymptomatic, but also because most individuals with symptomatic infections do not have access to, or do not seek clinical care, and those that do are typically not offered diagnostic testing. Testing is generally limited to individuals with severe symptoms or comorbidities. HuNoV is not a reportable disease in many countries, including the United States (US), so the data on the number of confirmed infections is not consistently recorded. Run by the US Center for Disease Control (CDC), Calicinet^2^ compiles information about HuNoV outbreaks and genomic sequences, and sentinel laboratories voluntarily submit HuNoV test positivity data to the National Respiratory and Enteric Virus Surveillance System (NREVSS)^3^ .

Indirect estimates suggest that annually, HuNoV causes between 19 and 21 million symptomatic illnesses and 570-800 deaths in the US^4^. HuNoV infections can be asymptomatic; a study conducted in England found 12% prevalence of asymptomatic HuNoV infections in the population^5^ while Teunis et al.^6^ report 32% of volunteers infected by HuNoV in a feeding study did not develop symptoms. There are two common genotypes of HuNoV: HuNoV GI and HuNoV GII. HuNoV GII.4 is responsible for the greatest proportion of infections globally^7^. Individuals with inactivated FUT2 enzyme (“non-secretors”) are resistant to infection by some HuNoV genotypes^6, 8^.

Wastewater-based epidemiology (WBE) is a low-bias approach for assessing disease burden in a community. Municipal wastewater contains human excretions including feces, urine, saliva, and mucus, that enter the wastewater systems via toilets, showers, sinks, and any other drains in buildings. All individuals using these drains in the sewershed contribute to wastewater. WBE has been used extensively during the COVID-19 pandemic to assess trends in SARS-CoV-2 infections^9^, and more recently, it has been used to understand occurrence and trends in Mpox virus^10^, influenza A^11, 12^, respiratory syncytial virus^13^ (RSV), human metapneumovirus, parainfluenza, seasonal coronaviruses, and rhinovirus infections^14^. Concentrations of nucleic-acids from these viruses correlate to clinical measures of disease occurrence, including incident case rates and test positivity rates, in the contributing populations. WBE has also provided insights into *Salmonella*^15^, hepatitis A^16^, and poliovirus^17^ circulation in communities. Therefore, WBE may be a useful tool for better understanding HuNoV disease occurrence.

HuNoV is shed in high concentrations in stool, where concentrations can be as high as 10^11^ copies of genomic RNA per gram of stool^18^. Fecal shedding is similar among asymptomatic and symptomatic infections^19^. HuNoV can also be detected in saliva of infected individuals^20^ with mean concentrations on the order of 10^3^ copies genomic RNA per ml saliva. Given that HuNoV is found in high concentrations in multiple excretions that contribute to wastewater, it is not surprising that studies have documented HuNoV in wastewater. Eftim et al.^21^ carried out a systematic review of HuNoV concentrations in wastewater globally and found that mean concentrations in liquid wastewater influent (globally) are 10^4.6^ copies/liter and tended to be highest in the winter and spring, consistent with the fact that HuNoV infections peak in the winter^22^. Moreover, concentrations of HuNoV GII were higher in wastewater influent than those of HuNoV GI, particularly in North America.

Raw wastewater is a complex mixture of human excretions, food waste, water, trash, and industrial inputs. It contains both a liquid and solid phase. A number of studies suggest that viral nucleic acids and viruses present in raw wastewater tend to adsorb to the solids in wastewater^10, 12, 23^. This has been shown to occur in both experimentally inoculated laboratory studies and in observations of raw wastewater for a wide range of enveloped and non-enveloped viruses^24^ . For example, Wolfe et al.^11^ showed that influenza A RNA was ∼1000 times high in the solids fraction compared to the liquid fraction of raw wastewater while Ye et al.^25^ showed in laboratory studies that when added to wastewater, enveloped viruses phi6 and murine hepatitis virus had a higher affinity to wastewater solids than non-enveloped MS2. Da Silva et al.^26^ showed that HuNoV RNA was associated with particles in waste stabilization pond influent.Based on this work there is reason to suspect HuNoV may also partition to wastewater solids, but to our knowledge, such data has not been reported in the literature.

Previous work has related HuNoV in wastewater to health. Huang et al.^27^ carried out a systematic review and meta-analysis of studies that linked information on HuNoV RNA in wastewater to the health of the contributing populations. They identified seven peer-reviewed studies that reported monthly concentrations of HuNoV RNA in wastewater influent and gastrointestinal infections (measured via a variety of metrics) in the contributing population; an eighth study ^28^ measured HuNoV RNA weekly in wastewater primary effluent serving approximately 10,000 people and compared their measurements to local gastroenteritis cases. These eight studies generally show good agreement between wastewater influent / primary effluent HuNoV RNA concentrations and measures of gastroenteritis in the communities, suggesting monitoring HuNoV in wastewater may provide useful insights into disease occurrence, and circulating genotypes, in communities. To date, however, no study has examined the occurrence of HuNoV RNA in wastewater solids, and investigated the possibility of using HuNoV RNA in solids for disease surveillance. Additionally, the HuNoV wastewater monitoring studies, to date, have measured HuNoV at a low frequency (primarily monthly). Receiving information on a more frequent basis (daily to weekly) may provide additional insights into disease occurrence and trends in populations.

The goal of the present study is to investigate the use of WBE using wastewater solids to understand HuNoV GII infection occurrence and trends. We focus on HuNoV GII because it has been shown to be more common than HuNoV GI in North America^21^. First, we conducted an experiment to examine the distribution of HuNoV GII RNA between the solid and liquid fractions of wastewater to inform use of the solid fraction for the study. As a first step to investigating the utility of WBE for understand HNoV GII disease occurrence, we first present data on concentrations of HuNoV GII genomic RNA in wastewater solids collected daily for four months at a large wastewater treatment plant (WWTP) that serves 1.5 million people, and compare the data to local clinical HuNoV positivity rates. We then present concentrations of HuNoV GII RNA in wastewater solids collected approximately three times per week from 145 WWTPs located in 26 states for up to 5 months, and compare to national estimates of clinical HuNoV positivity rates. Lastly, we derive a deterministic mass-balance model to estimate the number of individuals shedding HuNoV GII RNA given the concentrations measured in wastewater solids.

## Methods

This study was reviewed by the Institutional Review Board (IRB) at Stanford University and the IRB determined that this research does not involve human subjects as defined in 45 CFR 46.102(f) or 21 CFR 50.3(g).

### Daily sample collection at San José WWTP

San José (SJ) WWTP (San José-Santa Clara Regional Wastewater Facility) serves 75% (1,500,000 people) of Santa Clara County, California^29^. Daily samples of settled solids were collected between 15 November 2022 and 9 April 2023 (n=146). Daily samples of influent were collected between 28 February 2023 and 6 March 2023 (n = 7).

Fifty mL of settled solids were collected using sterile technique in clean bottles from the primary clarifier. Twenty-four hour composite samples were collected by combining grab samples every six hours. Samples were stored at 4°C, transported to the lab, and processed within six hours.

One-hundred mL of twenty-four hour composite influent samples were collected using sterile bottles. Samples were stored at 4°C, transported to the lab, and then stored for up to 7 days before analysis. Limited degradation of RNA targets over this period of time is expected^30, 31^.

### Three-day per week sample collection at national WWTPs

Samples were collected typically three times per week at up to 145 WWTPs between November 16, 2022 and April 9, 2023 (Figure S1 and Table S1). Samples of settled solids were collected from the primary clarifier, or solids were obtained from raw influent by either using an Imhof cone^32^, or allowing the influent to settle for 10-15 mins, and using a serological pipette to aspirate the settled solids into a falcon tube. Samples were collected by WWTP staff and sent at 4°C to our laboratory where they were processed immediately. The time between sample collection and receipt at the lab was typically between 1-3 days, during this time limited degradation of the RNA targets is expected^30, 31^. Table S1 provides additional information on the WWTPs including populations served and number and type of samples. In total, these WWTPs serve 10.9% of the US population. Note that the 145 WWTPs include San Jose WWTP. A total of 6911 samples were collected and analyze.

### Solid pre-analytical methods

Solids were first dewatered by centrifugation, and then dewatered solids were suspended in DNA/RNA Shield (Zymo Research, Irvine, CA) at a concentration of 0.75 mg (wet weight)/ml. The DNA/RNA shield was spiked with bovine coronavirus (BCoV) vaccine as a RNA recovery control. This concentration of solids was chosen as it was shown to alleviate inhibition in downstream RT-PCR^33^. An additional aliquot of dewatered solids was dried in an oven^29^ to determine its dry weight so that measured concentrations of nucleic acid targets could be normalized to gram dry weight. RNA was extracted from 10 or 6 (Table S1) replicate aliquots of dewatered settled solids suspended in the DNA/RNA Shield using the Chemagic Viral DNA/RNA 300 kit H96 for the Perkin Elmer Chemagic 360 (Perkin Elmer, Waltham, MA) followed by PCR inhibitor removal with the Zymo OneStep-96 PCR Inhibitor Removal kit (Zymo Research, Irvine, CA) The pre-analytical methods described here are provided in detail in other publications and on protocols.io^29, 33–36^.

### Influent pre-analytical methods

For each sample, 10 replicate aliquots were processed using an affinity-based capture method with magnetic hydrogel Nanotrap Particles with Enhancement Reagent 1 (Ceres Nanosciences, Manassas, VA) on 10 mL of sample to concentrate viral particles using a KingFisher Flex system following vendor instructions. RNA was then extracted from the each concentrated aliquot using the MagMAX Viral/Pathogen Nucleic Acid Isolation Kit (Applied Biosystems, Waltham, MA) on the KingFisher Flex platform to obtain purified nucleic acids which were then process through a Zymo OneStep-96 PCR Inhibitor Removal kit (Zymo Research, Irvine, CA).

### Digital droplet RT-PCR analytical methods

Each replicate RNA extract from each sample (10 or 6 per sample) was subsequently processed immediately to measure viral RNA concentrations using digital RT-PCR. We quantified the number of copies of the HuNoV GII ORF1-ORF2 junction using a previously established assay that has been shown to be highly specific to HuNoV GII^37^. Ten or 6 replicate wells (Table S1) were run for each sample. We also measured concentrations of pepper mild mottle virus (PMMoV) RNA; PMMoV is highly abundant in wastewater globally^38^ and is used as an internal recovery and fecal strength control^39^. We also measured concentrations of BCoV RNA. Ten or 2 replicate wells (Table S1) were run for each sample for PMMoV and BCoV in duplex). Extraction negative (BCoV spiked buffer, 3 wells) and positive (buffer spiked with positive control cDNA of targets, 1 well) controls, and PCR negative (molecular grade water, 3 wells) and positive controls (gene block cDNA, 1 well) were run on each 96 well plate. Primers and probes are provided in Table 1.

**Table 1.**
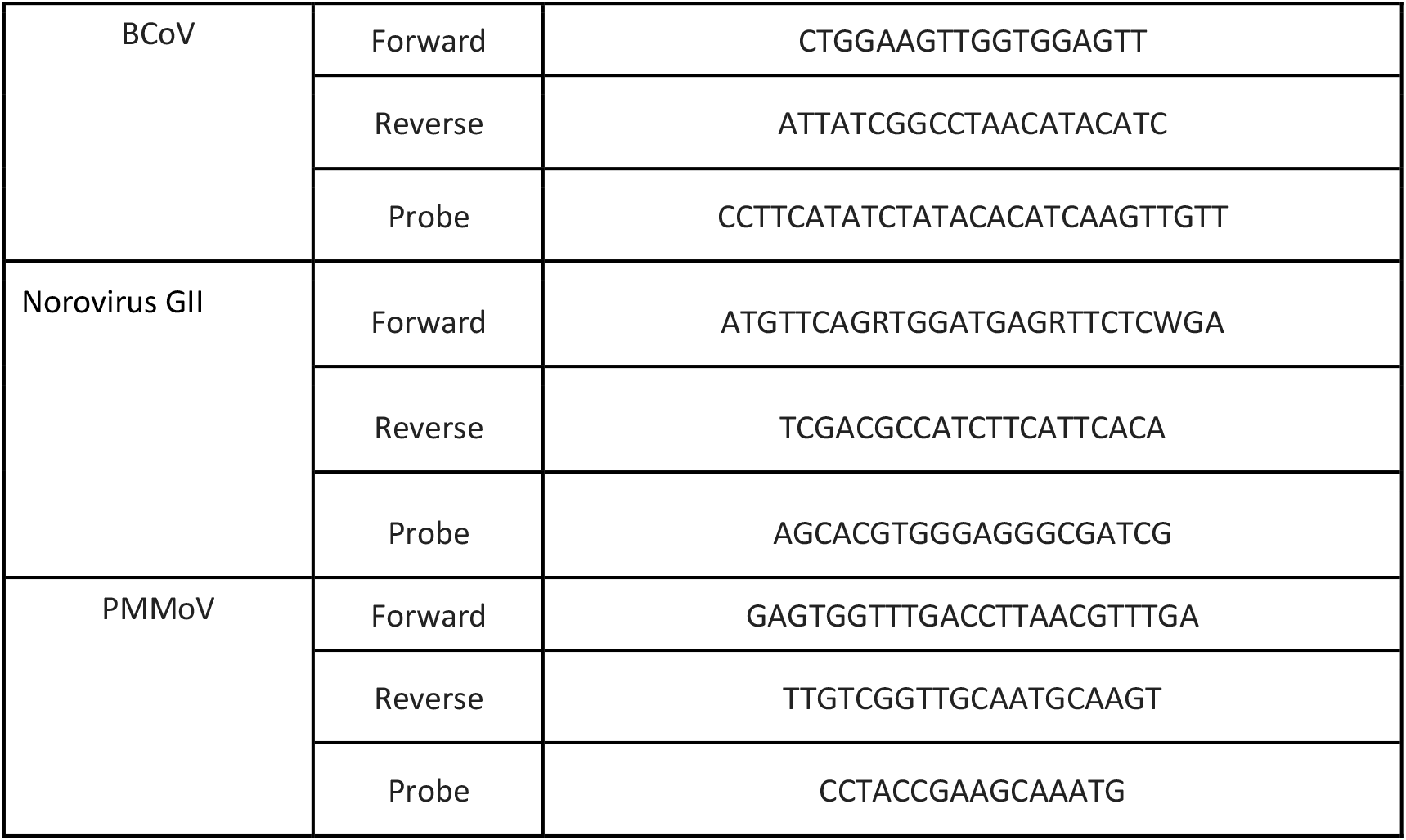
Forward and reverse primers, and probe sequences for detection of viral nucleic acids in this study. Primers and probes were purchased from Integrated DNA Technologies (IDT, Coralville, Iowa). All probes contained fluorescent molecules and quenchers (5ʹ FAM and/or HEX/ZEN/3ʹ IBFQ). FAM, 6-fluorescein amidite; HEX, hexachloro-fluorescein; ZEN, a proprietary internal quencher from IDT; IBFQ, Iowa Black FQ.

ddRT-PCR was performed on 20 µl samples from a 22 µl reaction volume, prepared using 5.5 µl template, mixed with 5.5 µl of One-Step RT-ddPCR Advanced Kit for Probes (Bio-Rad 1863021), 2.2 µl of 200 U/µl Reverse Transcriptase, 1.1 µl of 300 mM dithiothreitol (DDT) and primers and probes mixtures at a final concentration of 900 nM and 250 nM respectively. Primer and probes for assays were purchased from Integrated DNA Technologies (IDT, San Diego, CA) (Table 1). HuNoV was measured in reactions with undiluted template whereas PMMoV and BCoV assays were run in duplex on template diluted 1:100 in molecular grade water.

Droplets were generated using the AutoDG Automated Droplet Generator (Bio-Rad, Hercules, CA). PCR was performed using Mastercycler Pro (Eppendforf, Enfield, CT) with with the following cycling conditions: reverse transcription at 50°C for 60 minutes, enzyme activation at 95°C for 5 minutes, 40 cycles of denaturation at 95°C for 30 seconds and annealing and extension at 59°C (for HuNoV GII) or 56°C (for PMMoV/BCoV) for 30 seconds, enzyme deactivation at 98°C for 10 minutes then an indefinite hold at 4°C. The ramp rate for temperature changes were set to 2°C/second and the final hold at 4°C was performed for a minimum of 30 minutes to allow the droplets to stabilize. Droplets were analyzed using the QX200 or the QX600 Droplet Reader (Bio-Rad). A well had to have over 10,000 droplets for inclusion in the analysis. All liquid transfers were performed using the Agilent Bravo (Agilent Technologies, Santa Clara, CA).

Thresholding was done using QuantaSoft™ Analysis Pro Software (Bio-Rad, version 1.0.596). In order for a sample to be recorded as positive, it had to have at least 3 positive droplets. Replicate wells were merged for analysis of each sample.

Concentrations of RNA targets were converted to concentrations per dry weight of solids or per volume of influent (copies per gram dry weight (cp/g), or copies per ml (cp/ml), respectively) using dimensional analysis. The total error is reported as standard deviations and includes the errors associated with the Poisson distribution and the variability among the 10 replicates. Three positive droplets across 6 and 10 merged wells corresponds to a concentration between ∼300-600 cp/g and ∼500-1000 cp/g for solids, respectively, (the range in values is a result of the range in the equivalent mass of dry solids added to the wells) and three positive droplets across 10 merged wells corresponds to 1 cp/ml for influent.

### Inhibition testing

Inhibition of PMMoV and BCoV assays is not expected since RNA diluted 1:100 was used as template, and such high dilutions usually alleviate inhibition^40^. However, we tested for inhibition for the HuNoV GII assay in the influent and solids RNA extracts as RNA extracts were run neat as template in those reactions and inhibition could be present. Our previous work with our pre-analytical and analytical workflow suggests no RT-PCR inhibition^33^.

RNA extract from an influent sample (collected on 6 March 2023) was selected at random. It was tested for HuNoV GII following exactly the digital droplet RT-PCR analytical methods as follows: RNA extract was run neat as template, and then RNA extract was diluted 1:10 and 1:20 as template. The concentrations of HuNoV (in units of cp/ml) measured in the influent using the different diluted templates were compared to determine whether or not significant inhibition was present.

Two solids samples were chosen at random and rerun according to the preanalytical and analytical methods using the following concentrations of suspension of solids in DNA/RNA shield in the preanalytical processing: 7.5 mg/ml, 15 mg/ml, 37.5 mg/ml, and 75 mg/ml. The concentrations of HuNoV measured in the solids samples (units of cp/g) using the different mass concentration in the suspensions were compared to determine whether or not significant inhibition was present.

### HuNoV positivity rate data

There is no data on the incidence and prevalence of HuNoV infections in the United States. As such, we use HuNoV clinical test positivity rates to infer information about HuNoV infections. We used two sources of positivity rate data.

We used positivity rates from Stanford Health Care Clinical Microbiology Laboratory (hereafter “clinical laboratory”) for comparison to the wastewater data from SJ WWTP. The clinical laboratory tests stool specimens of symptomatic patients for HuNoV using the BIOFIRE GI panel (bioMérieux, Inc., Salt Lake City, UT). The clinical laboratory is among the largest in Santa Clara County, CA, where SJ WWTP is located. Although patients may or may not live in the service area of the WWTP, there is a high likelihood they do since the WWTP serves 75% of the county residents. We assumed that the HuNoV positivity rates recorded at the clinical laboratory are reasonable estimates for the positivity rates for residents in the sewershed.

We used national clinical HuNoV positivity rates publicly available through CDC NREVSS^3^. NREVSS compiles positivity rate data from sentinel laboratories across the country and reports positivity rates as 3-week, centered rolling means. Not every state participates in the voluntary reporting program, and not all sentinel laboratories provide HuNoV data to NREVSS; the exact states and laboratories reporting are not publicly available. We used the data as publicly available for the entire US for comparison to the wastewater data from WWTPs included in this study. It is assumed that the positivity rates for HuNoV described in this national data set is a reasonable estimate for the rates in populations served by the WWTPs.

### Statistical analyses

Data used in the study are not normally distributed, so we used non-parametric statistics to test hypotheses. We tested the null hypothesis that ratios of HuNoV RNA concentrations and PMMoV RNA in paired solids and liquids are the same using a Wilcoxon signed test. We tested the null hypothesis that the concentration of HuNoV RNA and HuNoV RNA / PMMoV RNA ratio are not associated with HuNoV positivity rates of clinical specimens collected at the clinical laboratory or compiled by CDC NVERSS using Kendall’s tau. Since positivity rates are aggregated by week, we used median wastewater concentrations for the same week as an independent variable. We computed a population-weighted national weighted average of HuNoV and HuNoV/PMMoV across all WWTPs for comparison to the national positivity rates (see SI). We used a p value of 0.05 to assess whether the null hypothesis should be rejected.

### Modeling

Previous work established a mass balance model relating the concentration of SARS-CoV-2 RNA in wastewater solids to the number of individuals excreting SARS-CoV-2 RNA into the wastewater^41^. We adopted that model to relate the concentration of HuNoV GII RNA in wastewater solids to the fraction of individuals excreting HuNoV into the wastewater as the mass balance model is not virus specific. The model relies on the assumption that fecal inputs into the wastestream are the main source of viral RNA, and that the fraction of the solids that are fecal in origin can be approximated using the concentration of PMMoV in the solids.

According to the model:

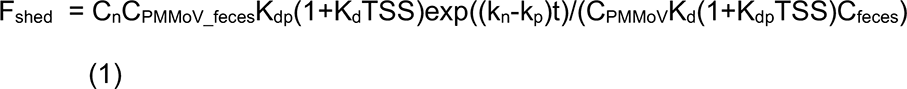

where C_n_ is the concentration of HuNoV RNA in wastewater solids, C_PMMoV_ is the concentration of PMMoV RNA in wastewater solids, C_PMMoV_feces_ is the concentration of PMMoV RNA in feces, k_p_ and K_dp_ are the PMMoV RNA first-order rate constant and partitioning coefficient, respectively, k_n_ and K_d_ are the HuNoV RNA first order decay constant and partitioning coefficient, respectively, t is the time sewage spends in the system, including the primary clarifier, prior to sampling, TSS is the total suspended solids in the influent of the treatment plant, C_feces_ is the concentration of HuNoV RNA shed in feces, and F_shed_ is the population fraction shedding HuNoV RNA in feces. The partitioning coefficient describes the ratio of the concentrations of HuNoV RNA in solids and liquids (unit of ml/g).

Given the small decay rate constants reported in the literature for both the HuNoV^30^ and PMMoV RNA targets ^31, 42^, and the short time t (< 1 day) that the material spends in the system between toilet and sampling point^41^, we neglect the decay term by assuming k_n_ and k_p_ are both small and that k_n_-k_p_∼0. Equation 1 can then be written:

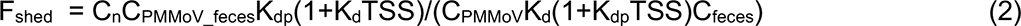

For the special case where K_d_ = K_dp_, equation (2) simplifies to

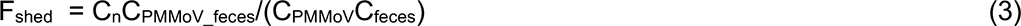

In applying the model, we assume viral RNA concentrations in settled solids from the primary clarifier are representative of concentrations associated with solids in wastewater, C_n_, and that samples represent a temporal composite of inputs.

Arts et al.^43^ provide a large dataset characterizing the distribution of C_PMMoV_feces_. Chan et al.^18^ provide data on HuNoV GII shedding in infected individuals within 48 hours of symptom onset; we digitized their data (from their figure 1A) using plot digitizer (https://plotdigitizer.com/) to characterize the distribution of C_feces._

**Figure 1.**
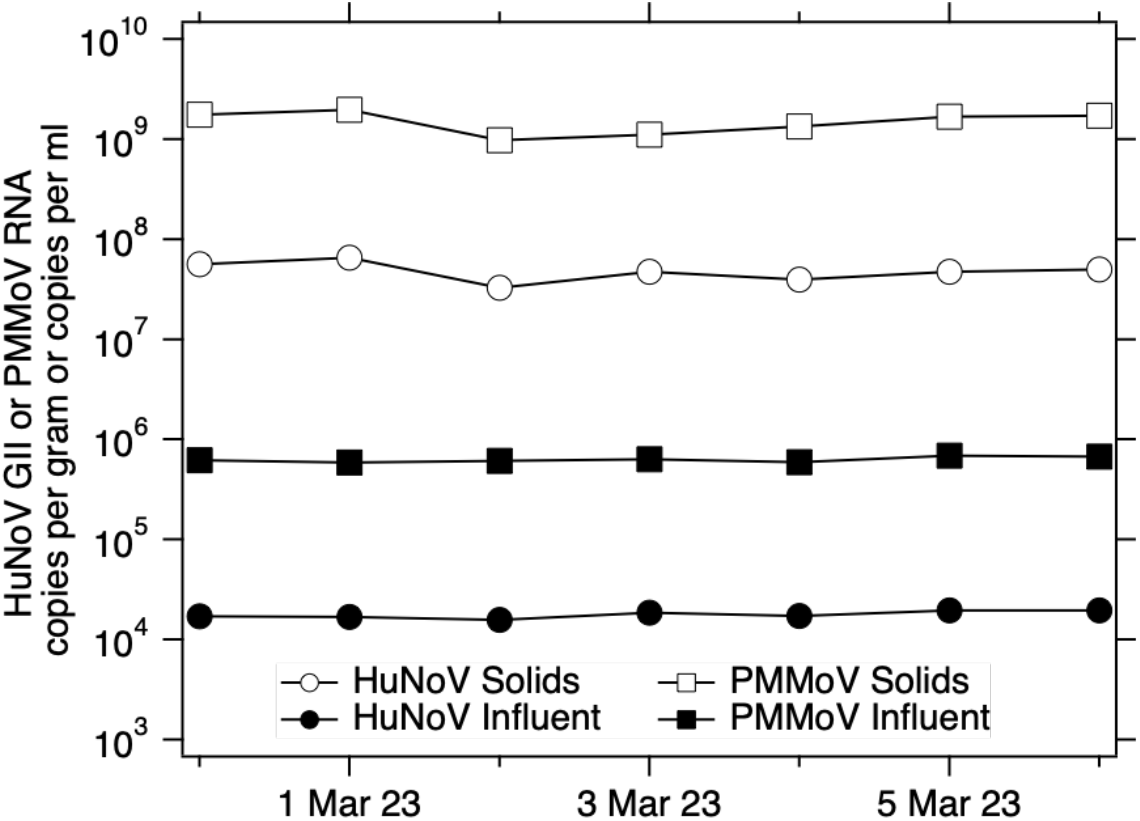
Concentrations of HuNoV GII and PMMoV RNA in influent and solids samples collected on the same day over seven consecutive days between 28 February 2023 and 6 March 2023. Each marker on the plot has an error bar, but the error bar is so small, it is covered by the marker. The error on the measurements is approximately 2-3%.

We modeled F_shed_ as a function of C_n_/C_PMMoV_ using a Monte Carlo simulation where values for C_feces_ and C_PMMoV_feces_ were drawn randomly from the distributions from Chan et al.^18^ and Arts et al.^43^, respectively. We varied C_n_/C_PMMoV_ as log_10_(C_n_/C_PMMoV_) from −4 to −1 in 0.1 increments. Ten thousand simulations were used to estimate median and interquartile ranges of F_shed_ for each C_n_/C_PMMoV_.

## Results

### Quality assurance and control (QA/QC)

Results are reported as suggested in the Environmental Microbiology Minimal Information guidelines^44^ (Figure S2). Extraction and PCR negative and positive controls performed as expected (negative and positive, respectively). We observed no evidence of inhibition of the HuNoV GII assay for solids or influent methods (Figure S3).

### HuNoV GII and PMMoV RNA in wastewater solids and influent

We measured concentrations of indigenous HuNoV GII RNA in matched wastewater influent and solids from SJ WWTP (Figure 1). Median BCoV recovery in these solids samples was 1.8 (n=7); values greater than 1 are likely a result of uncertainty in quantifying the amount of BCoV spiked in the DNA/RNA shield. We did not measure BCoV recovery in the influent samples, but according to the manufacturer, recovery of viral RNA from wastewater samples is typically 40-50%.

Matched samples were collected on the same day, but from different locations in the treatment train (influent from the inlet to the WWTP versus solids from the primary clarifier). HuNoV GII RNA concentrations in influent and solids were on the order of 10^4^ cp/ml and 10^7^ cp/g, respectively. The ratio of concentrations in matched solids and influent samples (K_d_) varied from 2100 to 3900 ml/g (median = 2500 ml/g, n = 7). Given the enrichment of HuNoV GII in solids relative to influent on an equivalent mass basis, we chose to measure HuNoV GII RNA in solids for our prospective study.

PMMoV RNA concentrations in influent and solids were on the order of 10^6^ cp/ml and 10^9^ cp/g, respectively. The ratio of concentrations in matched solids and influent samples (K_dp_) varied from 1600 to 3300 ml/g (median = 2400 ml/g, n = 7). K_dp_ and K_d_ were not different (Wilcoxon signed test, p = 0.54).

The ratio of HuNoV GII RNA concentrations and PMMoV RNA concentrations (HuNoV/PMMoV) ranged from 0.026 to 0.029 in influent, and 0.028 to 0.042 in solids. We tested the null hypothesis that HuNoV/PMMoV was not different in influent and solids and the null hypothesis was not rejected (Wilcoxon signed test, p=0.08).

### Prospective daily measurements of HuNoV GII RNA in wastewater solids at SJ WWTP and relation to clinical specimen testing

Median BCoV recovery across the 146 wastewater solids samples for SJ WWTP was 1.7. Recoveries higher than 1 are due to uncertainties associated with quantification of the amount of BCoV spiked into the DNA/RNA shield. No attempt was made to correct measurements by the BCoV recovery owing to complexities and uncertainties associated with measuring virus recovery^45^. Median PMMoV RNA concentrations across samples was 1.5×10^9^ cp/g, similar to measurements previously reported for the plant^14^. In addition, PMMoV levels were stable across samples (Figure S4, 25th percentile = 1.3×10^9^ cp/g, 75th percentile = 1.8×10^9^ cp/g) suggesting consistent fecal strength and RNA extraction efficiency across samples.

HuNoV GII RNA concentrations in wastewater solids varied from 4.5×10^6^ to 7.1 x 10^7^ cp/g with median concentrations of 2.3×10^6^ cp/g (Figure 2). Concentrations gradually increased throughout the study with occasional temporary drops in concentration. A similar trend is evident in the ratio of HuNoV GII RNA/ PMMoV RNA.

**Figure 2.**
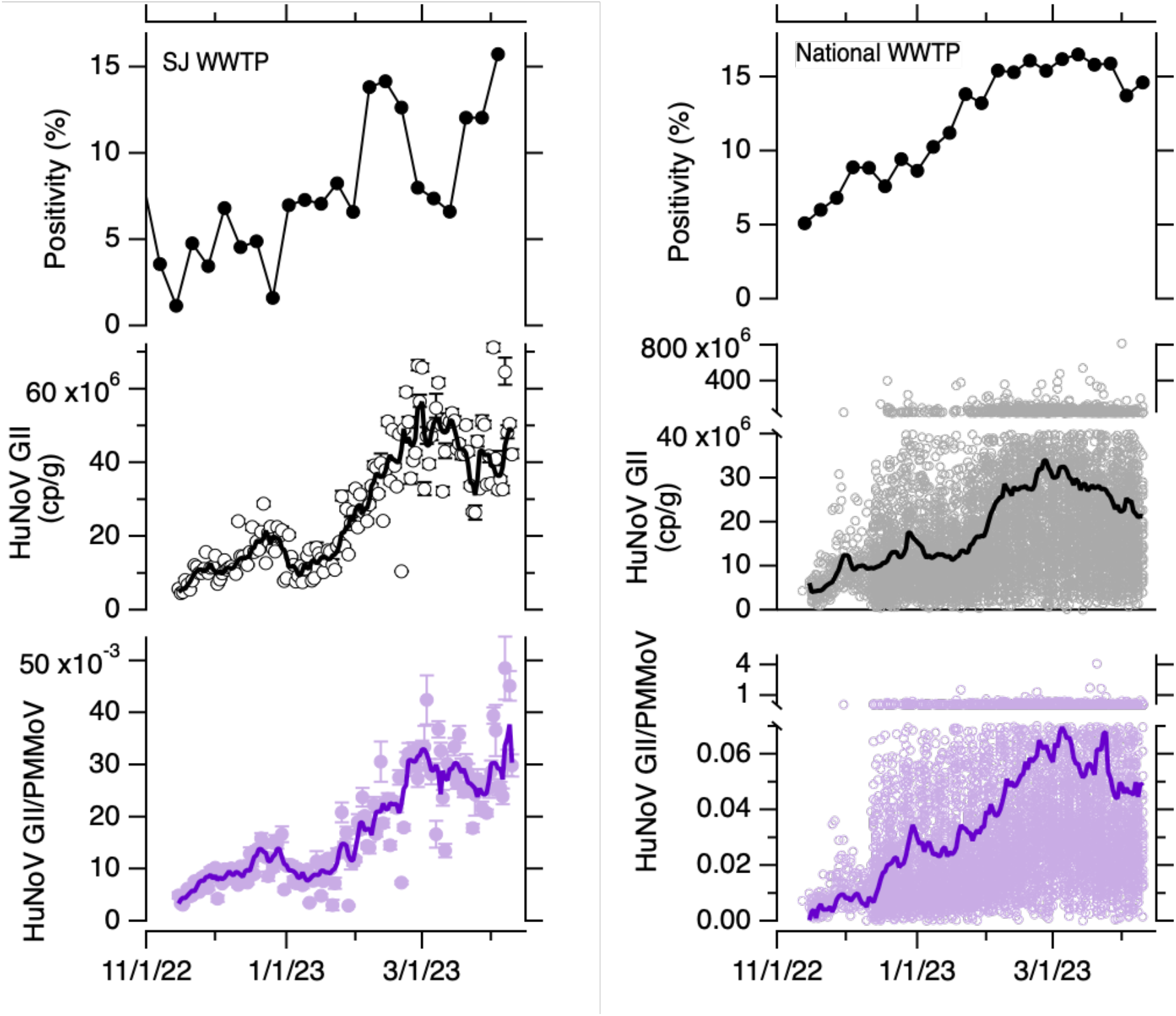
HuNoV clinical positivity rate and concentrations in wastewater solids. Top panels: Positivity rate for clinical specimens processed by the clinical laboratory (left) and by the sentinel laboratories participating in NVERRS (right). Middle panels and bottom panel: Concentrations of HuNoV GII RNA and HuNoV GII RNA normalized by PMMoV RNA, respectively. For the SJ WWTP, errors are provided a standard deviations as reported by the analysis software that incorporate variability among 10 replicates and Poisson error; if error bars are not visible, it is because they are smaller than the size of the marker. The error on the ratio is propagated from those of the values in the numerator and denominator. The 5-d trimmed average is shown as well as raw data as a solid line. For the national WWTP, raw data are shown for every WWTP, broken axes are needed to show higher measurements. The population-weighted average lines are shown as solid lines.

The clinical laboratory tested between 62 and 120 (median = 88) specimens per week between 14 November 2022 and 9 April 2023. The weekly HuNoV positivity rate varied from 1% to 16% (median = 7%). The weekly clinical positivity rate was positivity correlated to the weekly median HuNoV GII RNA concentrations in settled solids at the SJ WWTP (Kendall’s tau = 0.47, p = 0.003); the correlation was similar when median weekly HuNoV GII/PMMoV was used for the wastewater variable (Kendall’s tau= 0.45, p = 0.004).

### Prospective measurements of HuNoV GII RNA in wastewater solids at 145 WWTPs nationally and relation to clinical specimen testing

Median BCoV recovery across all the samples (n=8410) was 1.2. Median PMMoV RNA concentrations across samples was 5.4×10^8^ (25th percentile = 3×10^8^, 75th percentile = 10×10^8^ cp/g). HuNoV GII RNA concentrations in wastewater solids varied from 0 to 8.2×10^8^ cp/g with median concentrations of 1.5×10^7^ cp/g (Figure 2). The population-weighted average HuNoV GII RNA concentration across all the WWTP (Figure 2) increased throughout the study period and began to decrease at the end of the time series.

Nationally, weekly-aggregated clinical specimen positivity rates from NVERSS varied from 5% to 17%. The median weekly population weighted average HuNoV GII RNA was positively correlated to the NVERSS positivity rate date (Kendall’s tau = 0.74, p<10^-7^). The correlation between HuNoV GII RNA/PMMoV RNA and positivity rate was similar (Kendall’s tau = 0.76, p<10^-7^).

### Model Results

The distributions C_PMMoV_feces_ and C_feces_ RNA in units of copies per gram are characterized by log_10_-means (log_10_-standard deviations) of 7.72 (1.71) and 8.39 (1.57), respectively. On examination of a Q-Q plot, the distributions are reasonably modeled as log-normal distributions.

The relationship between C_n_/C_PMMoV_ and F_shed_ is linear (equation 3, Figure S5). At SJ WWTP, C_n_/C_PMMoV_ varied between 0.003 and 0.05 during the study. Based on the model, these minimum and maximum wastewater solids concentrations correspond to F_shed_ (expressed as percent of individuals shedding) of 0.06% (median model output, 25th percentile = 0.002%, 75th percentile = 2%) and 1% (median model output, 25th percentile = 0.03%, 75th percentile = 35%). The daily estimates of F_shed_ are displayed in Figure 3.

**Figure 3.**
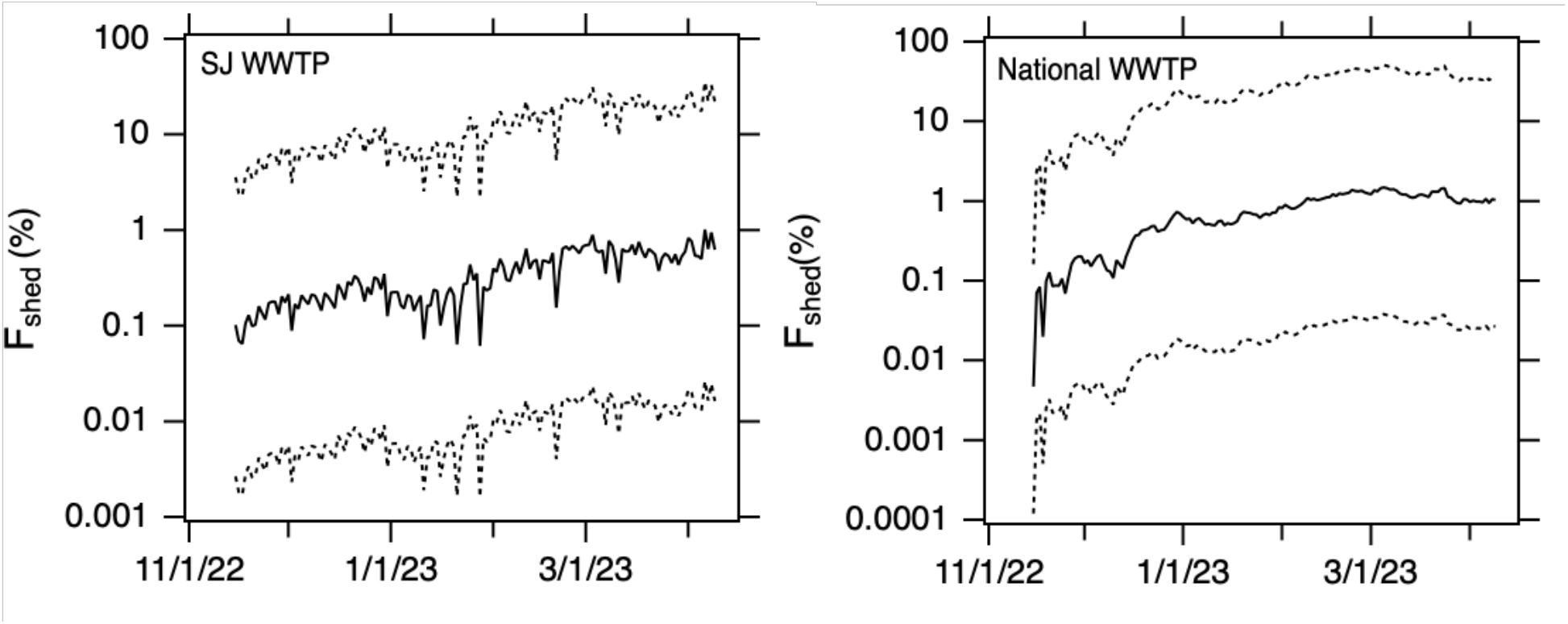
Modeled F_shed_ for SJ WWTP an the national WWTPs. Solid line is the median model output, and the 25th and 75th percentile model outputs are shown as dotted lines.

The population-weighted average C_n_/C_PMMoV_ across all 145 WWTP included in this study ranged from 0.0002 to 0.07. Model output for F_shed_, using the population-weighted average C_n_/C_PMMoV_ ranges from 0.004% (median model output, 25th percentile = 0.0001%, 75th percentile = 0.2%) to 1.5% (median model output, 25th percentile = 0.04%, 75th percentile =51%). The daily estimate of F_shed_ is displayed in Figure 3.

## Discussion

HuNoV GII RNA concentrations in wastewater solids are positively associated with clinical specimen positivity rates both at a single WWTP, and aggregated across WWTPs spanning the entire United States. This finding supports previous work that illustrates HuNoV RNA concentrations in liquid wastewater reflect HuNoV infections in the contributing communities, as summarized in the systematic review by Huang et al.^27^. Whereas most previous studies have studied co-variation in clinical measures of HuNoV infections and wastewater concentrations on monthly scales over the course of a year or more ^27^, the present work shows the relationship between wastewater solids HuNoV GII RNA concentrations and clinical measures occurrence co-vary over finer and shorter time scales. Wastewater measurements are available as quickly as 24 hours after a sample is received at a laboratory, so this work suggests that wastewater testing may provide real-time information on changes in disease occurrence for use by public health officials, clinicians, and the public by arriving in databases before data on clinical infections.

There are limitations associated with the clinical positivity rate data used in this study. It is generated using specimens from patients who may not live in the sewersheds. The specimens collected by the clinical laboratory are from individuals with severe symptomatic illness who sought medical care and the clinicians determined diagnostic testing was needed. Many of these patients likely had comorbidities. The data obtained from CDC NVERSS database are from sentinel laboratories located throughout the US, not necessarily in the states where wastewater data were collected; a list of the precise sentinel laboratories submitting HuNoV data, and the frequency at which they submit data is not publicly available. Therefore, the positivity rates do not necessarily reflect actual community HuNoV infection occurrence in the communities contributed to the wastewater system. Nevertheless, the agreement between the positivity rate and wastewater data lends credence to the wastewater data being reflective of community disease occurrence.

We found HuNoV GII RNA was three orders of magnitude higher in concentration in wastewater solids compared to liquid influent on a mass equivalent basis. We found PMMoV RNA was also concentrated in the solids relative to the liquid influent to a similar extent. This supports previous work that suggests viral nucleic acids partition to the solid phase of wastewater^10–12, 23–25^. Solids naturally concentrate the viral nucleic acids in the waste stream and can be used as a basis for analysis without the need for time-consuming pre-analytical processes that are not automatable^29^. Because PMMoV RNA and HuNoV GII RNA partition to wastewater solids to similar extents, their ratio in solids and liquid is not different. Additional research is warranted to understand if measurements in wastewater solids and influent can be readily compared after each is normalized to PMMoV concentrations.

We derived a model for relating HuNoV GII RNA concentrations in wastewater solids to the number of individuals in the community shedding HuNoV in feces. Owing to the fact that the partitioning of HuNoV GII and PMMoV RNA was not different, the model simplified significantly resulting in a linear relationship between the fraction of individuals in the sewershed shedding HuNoV GII RNA (F_shed_) and the ratio HuNoV GII RNA/PMMoV RNA in solids. HuNoV shedding drops off exponentially with time since day of infection^19^, so new infections likely contribute the most to HuNoV GII RNA measured in wastewater, as has also been suspected for SARS-CoV-2^41^. Therefore, F_shed_ likely approximates incident HuNoV infections in the contributing population rather than the overall prevalence of infection. Our model used measurements of shedding within the first 48 hours after symptom onset from Chan et al.^18^ and thus likely represents peak shedding^19^. Individuals shedding higher concentrations of virus in their stool (“super shedders”) will contribute the most to wastewater HuNoV GII RNA.

Summing daily F_shed_, estimated from the national data over the duration of the approximate five month study at the median and 75th percentile model outputs suggest that the entire contributing population was infected at least once with HuNoV during the study period. These estimates are probably unrealistic.On the other hand, summing the 25th percentile F_shed_ over the duration of the five month study suggests 3% of the contribution population was infected with HuNoV GII. This is consistent with indirect estimates that approximately 7% of the US population is infected with symptomatic HuNoV annually^4^. Assuming asymptomatic infections make up 30% of the infections^6^, our model at the 25th percentile estimates 2% of the population had a symptomatic infection over 5 months or, scaling linearly, 5% over 12 months. The 25th percentile model output for F_shed_ is calculated using higher C_n_ and gives greater weight to “super shedders”. Future efforts are needed to further refine models for estimating incidence and prevalence of disease from wastewater, and fully explore their usefulness. While these results align realistically with other estimates of disease occurrence, the large range in observed peak shedding for HuNoV^19^ demonstrates how difficult it may be to model incidence or prevalence of the disease with precision.

WBE can provide information on HuNoV infections in communities and has also been shown to be useful for identifying circulating HuNoV genotypes^46^. It should be noted that WBE should not replace individual testing, as those are used to inform clinical decision making for severely ill patients. However, knowledge gleaned from wastewater may be used by public health professionals to make recommendations on hand washing, surface disinfection, or other behaviors to reduce transmission of HuNoV, or medical doctors to inform clinical decision making. Additional molecular epidemiology applications to HuNoV in wastewater may provide insight into viral genomic diversity and evolution, as well as vaccine development efforts. It should be noted that our study was limited to HuNoV GII, but HuNoV GI likely also contributes to HuNoV disease occurrence.

## Data availability statement

All wastewater data used in this study are available through the Stanford Digital Repository at https://doi.org/10.25740/rk281xb8780.

## Acknowledgements

We acknowledge the numerous people across the country who contributed to wastewater sample collection, Payal Sakar at SJ WWTP for her support and contribution to the work, and Allegra Koch for her support with the literature review. This research was performed on the ancestral and unceded lands of the Muwekma Ohlone people. We pay our respects to them and their Elders, past and present, and are grateful for the opportunity to live and work here.

## Author contributions

A. B. Boehm: Conceptualization, Methodology, Validation, Formal analysis, Investigation, Resources, Data Curation, Writing - Original Draft, Writing - Review & Editing, Visualization, Supervision, Project administration, Funding acquisition

M. K. Wolfe: Conceptualization, Methodology, Data Curation, Validation, Writing - Review & Editing

B. Hughes: Methodology, Validation, Investigation, Data Curation, Writing - Review & Editing

D. Duong: Methodology, Validation, Investigation, Data Curation, Writing - Review & Editing

A. Bidwell: Data Curation, Writing - Original Draft, Writing - Review & Editing, Visualization Bradley White: Methodology, Validation, Investigation, Resources, Data Curation, Writing - Review & Editing, Supervision

N. Banaei: Data Curation, Writing - Review & Editing

## Notes

### Competing Interest Statement

Bradley White, Dorothea Duong, and Bridgette Hughes are employees of Verily Life Sciences. The other authors have no competing interests.

### Funding Statement

This work was supported by gifts from the CDC Foundation and the Sergey Brin Family Foundation.

