## Supporting Information for "Human norovirus (HuNoV) GII RNA in wastewater solids at 145 United States wastewater treatment plants: Comparison to positivity rates of clinical specimens and modeled estimates of HuNoV GII shedders"

for

### Calculation of population weighted average HuNoV and HuNoV/PMMoV

The population weighted average HuNoV and HuNoV/PMMoV is calculated using the raw data for each site. For each date, an individual WWTP, the value measured for the day is multiplied by the WWTP's population served, as reported by the WWTP. These products and then summed across each day, and divided by the total population of all WWTPs. This calculation results in the population weighted average for day d, as in the following equation for the 145 selected plants:

$$Population\ weighted\ average\ of\ Z(d)\ = \left(\sum_{n=1}^{145} pop_n * Z(d)\right) / \left(\sum_{n=1}^{145} pop_n\right)$$

Where Z(d) is either HuNoV or HuNoV/PMMoV on day d, $pop_n$ is the population served by WWTP n,and n is a counter that goes from 1 to 145 and covers each WWTP. Prior to calculating the aggregated line, non-detect values are set to 500 cp/g (approximately the limit of detection for HuNoV), and for Z = HuNoV/PMMoV, 500 cp/g was normalized by the average PMMoV value for the WWTP. For days when a WWTP did not have a sample, Z was estimates using linear interpolation between the previous and next values with respect to d.

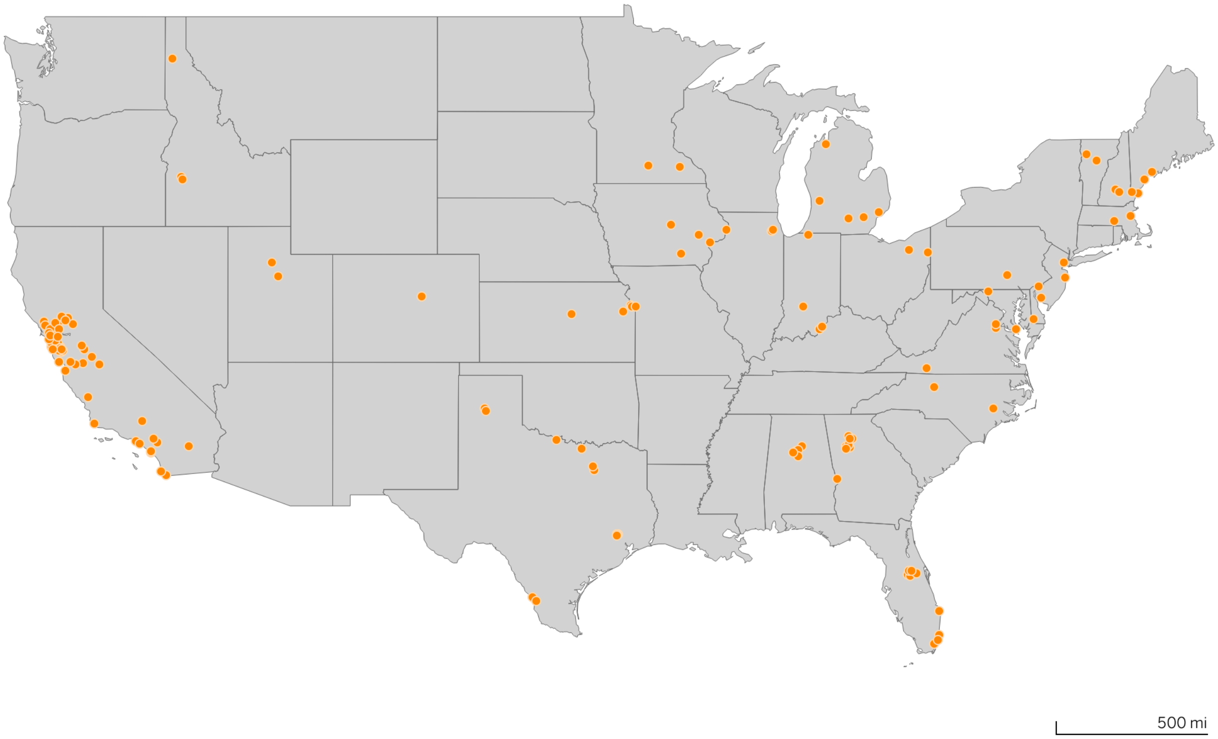


Figure S1. Map of 145  WWTPs. Orange dot indicates WWTP location.

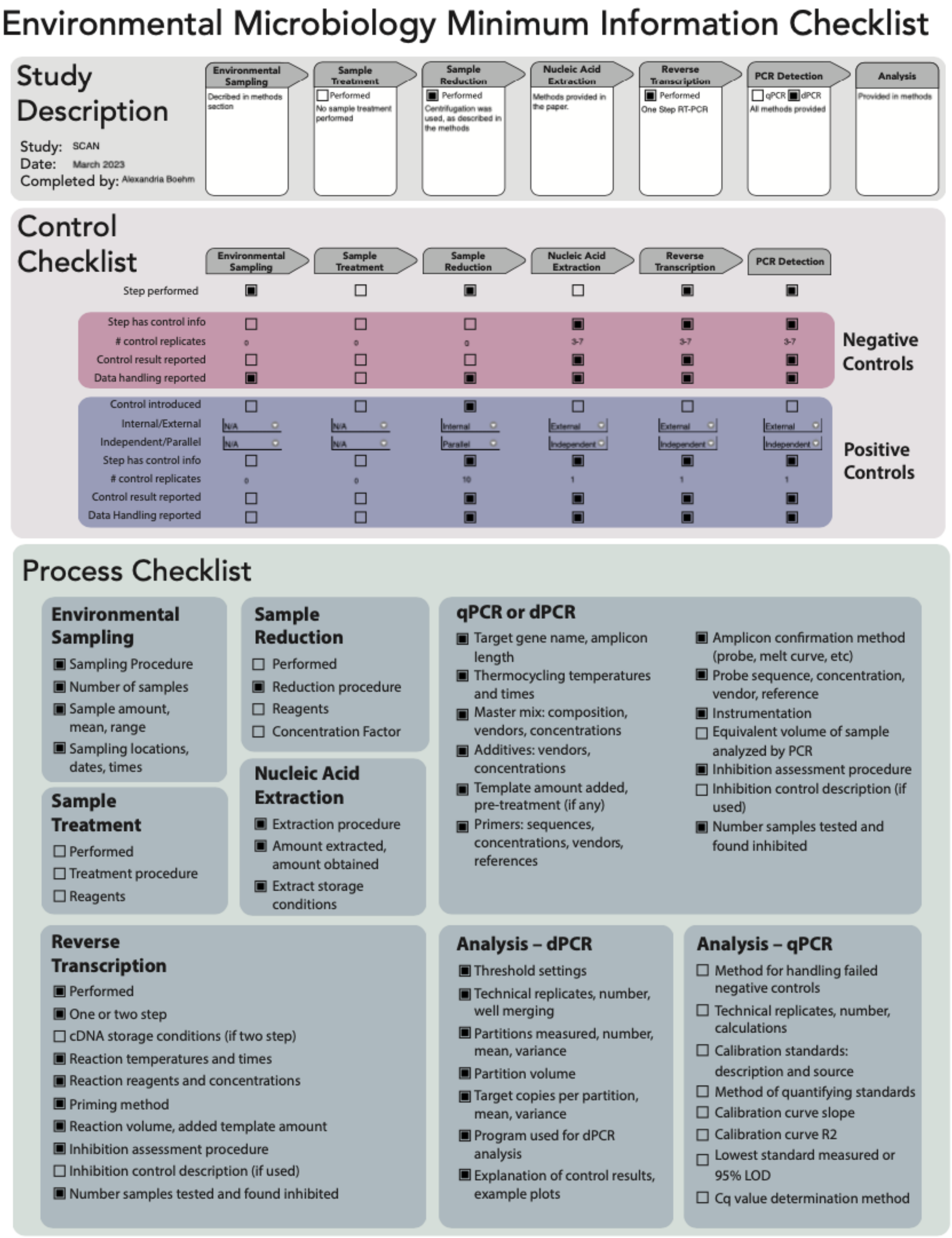


Figure S2. EMMI checklist from Borchardt et al.[1]

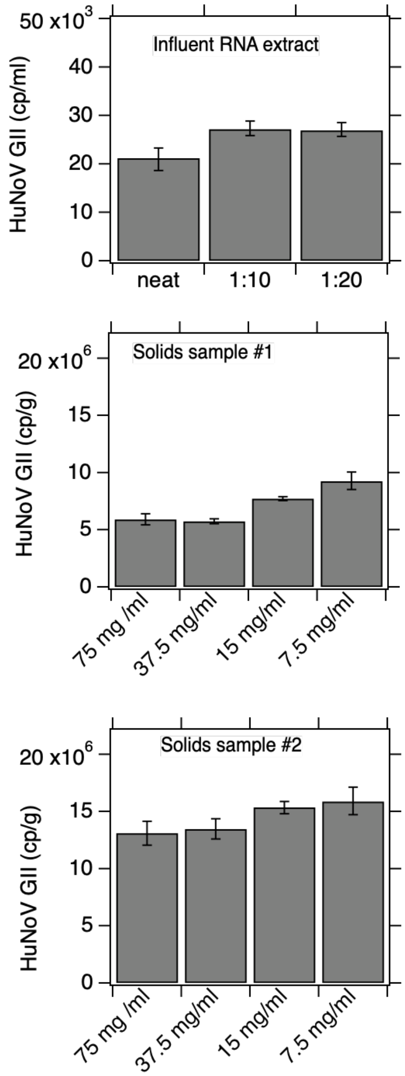


Figure S3. Data from inhibition testing. Concentration of HuNoV GII RNA is shown on the y-axis in all panels. Top panel. Concentrations derived using different dilutions of RNA extract from wastewater influent samples as template in the dd-RT-PCR; neat indicates not diluted. Similar results (less than 2X differences)[2] across dilutions indicate lack of evidence of inhibition. Middle and bottom panels. Results for concentrations in wastewater solids obtained using different mass concentrations of solids in DNA/RNA shield. Similar concentrations (less than 2X differences)[2] as the concentration is decreased suggests absence of inhibition.

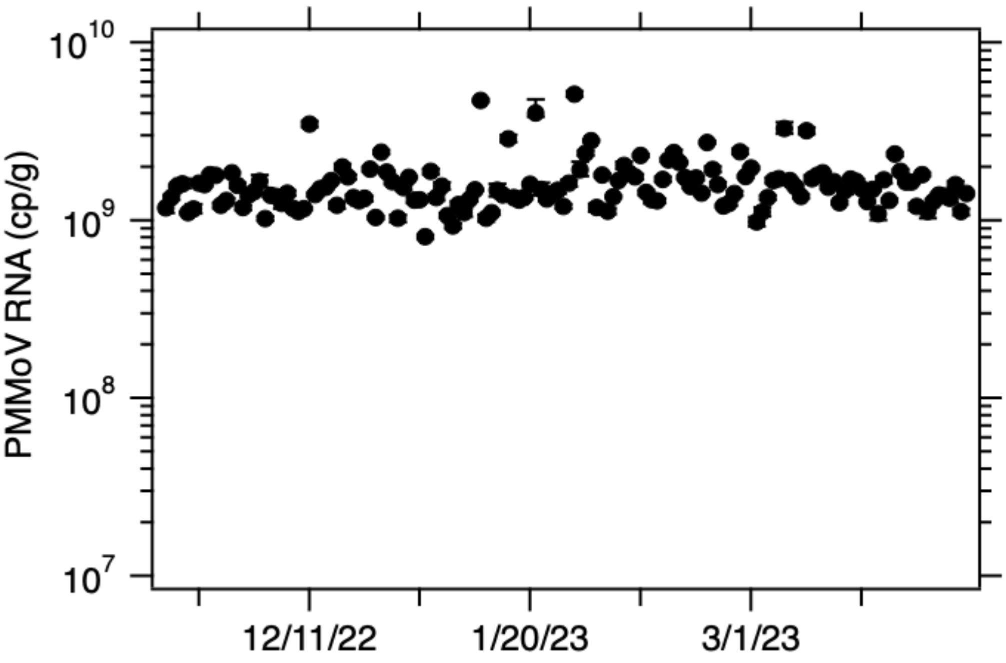


Figure S4. Concentrations of PMMoV RNA in wastewater solids from SJ WWTP. Errors are standard deviations. If the error bar cannot be seen, then it is smaller than the marker.

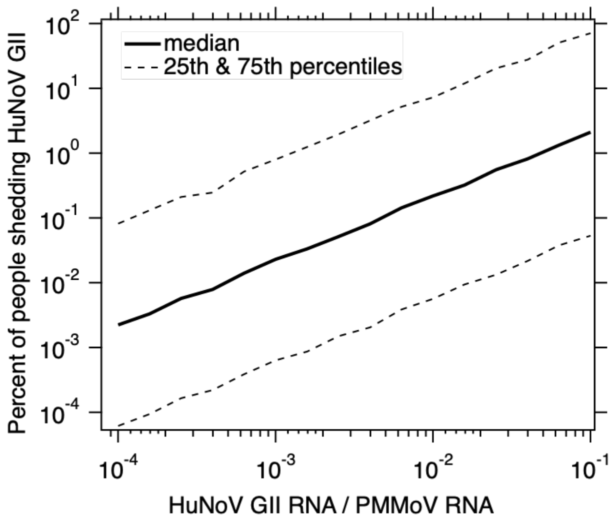

Figure S5. Modeled percent of individuals shedding HuNoV GII in their feces ($F_{shed}$) as a function of the ratio of HuNoV GII RNA and PMMoV RNA in wastewater solids.

Table S1: Details of the 145 WWTPs included in the study. The state where the WWTP is located, additional details regarding plant location, the sample type provided, the sample county and the reported population served. If an * appears next to the name (8 WWTPs total in CA), then 10 replicates were used to measuring HuNoV, PMMoV and BCov. Otherwise, 6 replicates were used to measure HuNoV and 2 to measure PMMoV and BCoV.

| State | Plant | Sample Type | Sample Count | Population Served |
|---|---|---|---|---|
| Alabama | Bessemer, AL | Liquids | 50 | 225,000 |
| | Cahaba River, Birmingham, AL | Liquids | 34 | 95,000 |
| | Fultondale, AL | Liquids | 26 | 77,000 |
| | Pinson, AL | Liquids | 29 | 30,000 |
| | Village Creek, Birmingham, AL | Liquids | 50 | 200,000 |
| California | Coastal, Laguna Niguel, CA | Liquids | 47 | 48,000 |
| | Contra Costa County, CA | Solids | 42 | 484,800 |
| | Davis, CA | Solids | 88 | 68,000 |
| | Esparto, CA | Liquids | 46 | 4,006 |
| | Fairfield, CA | Solids | 50 | 155,000 |
| | Fremont, CA | Liquids | 31 | 229,476 |
| | Gilroy, CA* | Solids (Settling w Imhoof cones onsite) | 145 | 110,338 |
| | Half Moon Bay, CA | Liquids | 38 | 28,000 |
| | Hollister, CA | Liquids | 47 | 42,000 |
| | Indio, CA | Solids | 50 | 91,765 |
| | JB Latham, Laguna Niguel, CA | Liquids | 47 | 120,000 |

| State | Plant | Sample Type | Sample Count | Population Served |
|---|---|---|---|---|
| | Lancaster, CA | Liquids | 49 | 200,000 |
| | Las Gallinas, San Rafael, CA | Liquids | 50 | 30,000 |
| | Lompoc, CA | Liquids | 50 | 69,290 |
| | Los Angeles County, CA | Liquids | 51 | 3,500,000 |
| | Los Angeles, CA | Liquids | 51 | 4,000,000 |
| | Los Banos, CA | Liquids | 52 | 42,000 |
| | Madera, CA | Solids | 15 | 67,944 |
| | Marina, CA | Liquids | 44 | 262,000 |
| | Merced, CA | Solids (Settling w Imhoof cones onsite) | 50 | 91,000 |
| | Mill Valley, CA | Solids | 51 | 30,000 |
| | Modesto, CA | Solids | 50 | 230,000 |
| | Napa, CA | Solids | 46 | 83,300 |
| | Newark, CA | Liquids | 32 | 47,229 |
| | Novato, CA | Liquids | 50 | 53,000 |
| | Oakland, CA | Solids | 50 | 740,000 |
| | Oceanside, San Francisco, CA* | Solids | 140 | 250,000 |
| | Ontario, CA | Solids | 47 | 890,000 |
| | Pacifica, CA | Liquids | 49 | 40,000 |
| | Palo Alto, CA* | Solids | 145 | 236,000 |
| | Paso Robles, CA | Solids | 49 | 31,037 |

| State | Plant | Sample Type | Sample Count | Population Served |
|---|---|---|---|---|
| | Petaluma, CA | Liquids | 50 | 65,000 |
| | Redwood City, CA* | Solids | 145 | 199,000 |
| | Regional, Laguna Niguel, CA | Liquids | 46 | 129,000 |
| | Riverside, CA | Liquids | 32 | 350,000 |
| | Sacramento, CA* | Solids | 145 | 1,480,000 |
| | San Diego, CA | Liquids | 49 | 2,200,000 |
| | San Jose, CA* | Solids | 145 | 1,500,000 |
| | San Leandro, CA | Liquids | 50 | 50,000 |
| | San Mateo, CA | Solids | 46 | 150,000 |
| | San Rafael, CA | Liquids | 30 | 104,250 |
| | Santa Cruz County, CA | Solids (Settling w Imhoof cones onsite) | 49 | 160,000 |
| | Santa Cruz, CA | Solids (Settling w Imhoof cones onsite) | 49 | 160,000 |
| | Santa Rosa, CA | Solids (Settling w Imhoof cones onsite) | 47 | 230,000 |
| | Sausalito, CA | Liquids | 35 | 18,000 |
| | South San Diego, CA | Solids | 36 | 1,600,000 |
| | Southeast San Francisco, CA* | Solids | 142 | 750,000 |
| | Sunnyvale, CA* | Solids | 144 | 153,000 |
| | Turlock, CA | Liquids | 50 | 86,000 |
| | Union City, CA | Liquids | 31 | 68,150 |

| State | Plant | Sample Type | Sample Count | Population Served |
|---|---|---|---|---|
| | Vallejo, CA | Liquids | 48 | 121,000 |
| | West Contra Costa County, CA | Liquids | 43 | 100,000 |
| | West Railroad, San Rafael, CA | Liquids | 31 | 25,000 |
| | Windsor, CA | Liquids | 43 | 28,000 |
| | Winters, CA | Liquids | 46 | 7,286 |
| | Woodland, CA | Liquids | 50 | 59,000 |
| Colorado | North, Parker, CO | Liquids | 47 | 35,000 |
| | South, Parker, CO | Liquids | 48 | 25,000 |
| Delaware | Seaford, DE | Solids | 26 | 13,172 |
| Florida | Altamonte Springs, FL | Liquids | 44 | 95,000 |
| | Eastern, Orange County, FL | Solids (Settling w Imhoof cones onsite) | 45 | 195,299 |
| | Jupiter, FL | Liquids | 50 | 90,000 |
| | Key Biscayne, FL | Liquids | 28 | 829,725 |
| | North Miami, FL | Liquids | 35 | 776,150 |
| | Northwest, Orange County, FL | Solids (Settling w Imhoof cones onsite) | 47 | 66,690 |
| | South Miami, FL | Liquids | 35 | 920,528 |
| | South, Orange County, FL | Solids (Settling w Imhoof cones onsite) | 45 | 183,009 |
| | Southwest, Orange County, FL | Liquids | 44 | 50,000 |
| Georgia | Big Creek, Roswell, GA | Liquids | 50 | 189,593 |
| | College Park, GA | Liquids | 50 | 73,821 |

| State | Plant | Sample Type | Sample Count | Population Served |
|---|---|---|---|---|
| | Columbus, GA | Solids | 33 | 278,000 |
| | Johns Creek, Roswell, GA | Liquids | 50 | 84,486 |
| | Little River, Roswell, GA | Liquids | 50 | 12,818 |
| | RM Clayton, Atlanta, GA | Liquids | 48 | 94,000 |
| | South River, Atlanta, GA | Liquids | 48 | 94,000 |
| | Utoy Creek, Atlanta, GA | Liquids | 48 | 94,000 |
| Idaho | Coeur d'Alene, ID | Solids | 48 | 50,540 |
| | Lander Street, Boise, ID | Liquids | 36 | 108,556 |
| | West Boise, ID | Liquids | 36 | 186,901 |
| Illinois | Glen Ellyn, IL | Solids (Settling w Imhoof cones onsite) | 42 | 86,000 |
| | Wheaton, IL | Solids (Settling w Imhoof cones onsite) | 50 | 63,000 |
| Indiana | Bloomington, IN | Liquids | 44 | 56,090 |
| | Downtown, Jeffersonville, IN | Liquids | 46 | 25,000 |
| | North, Jeffersonville, IN | Liquids | 45 | 25,000 |
| | South Bend, IN | Liquids | 38 | 130,000 |
| Iowa | Clinton, IA | Solids | 36 | 29,300 |
| | Coralville, IA | Liquids | 31 | 23,000 |
| | Marshalltown, IA | Liquids | 33 | 27,400 |
| | Muscatine, IA | Solids | 50 | 24,400 |
| | Ottumwa, IA | Liquids | 49 | 25,529 |

| State | Plant | Sample Type | Sample Count | Population Served |
|---|---|---|---|---|
| Kansas | Kaw Point, Kansas City, KS | Solids (Settling w Imhoof cones onsite) | 39 | 90,000 |
| | Lawrence, KS | Solids (Settling w Imhoof cones onsite) | 44 | 80,000 |
| | P20, Kansas City, KS | Solids (Settling w Imhoof cones onsite) | 39 | 35,000 |
| | Salina, KS | Solids | 50 | 47,000 |
| | Wolcott, Kansas City, KS | Liquids | 38 | 15,000 |
| Kentucky | Louisville, KY | Solids (Settling w Imhoof cones onsite) | 14 | 423,913 |
| Maine | Brunswick, ME | Liquids | 39 | 10,000 |
| | Portland, ME | Liquids | 36 | 65,000 |
| | York, ME | Liquids | 32 | 10,000 |
| Maryland | Hagerstown, MD | Liquids | 45 | 90,000 |
| | Hollywood, MD | Liquids | 33 | 55,000 |
| Massachusetts | Boston, MA | Solids | 48 | 2,400,000 |
| | Millbury, MA | Liquids | 18 | 250,000 |
| Michigan | Ann Arbor, MI | Liquids | 50 | 125,000 |
| | Jackson, MI | Solids | 51 | 90,000 |
| | Jenison, MI | Solids | 51 | 75,000 |
| | Traverse City, MI | Liquids | 33 | 30,623 |
| | Warren, MI | Liquids | 46 | 140,000 |
| Minnesota | Mankato, MN | Solids (Settling w Imhoof cones onsite) | 48 | 70,000 |
| | Rochester, MN | Solids (Settling w Imhoof cones onsite) | 49 | 120,000 |

| State | Plant | Sample Type | Sample Count | Population Served |
|---|---|---|---|---|
| New Hampshire | Dover, NH | Liquids | 41 | 30,000 |
| | Hall Street, Concord, NH | Liquids | 49 | 45,000 |
| | Penacook, Concord, NH | Liquids | 48 | 4,000 |
| New Jersey | Belmar, NJ | Solids | 50 | 52,672 |
| | Bridgeton, NJ | Liquids | 11 | 50,000 |
| | Newark, NJ | Solids | 43 | 1,500,000 |
| North Carolina | Kinston, NC | Liquids | 49 | 25,000 |
| | Winston-Salem, NC | Liquids | 42 | 92,000 |
| Ohio | Akron, OH | Solids | 35 | 365,000 |
| | Youngstown, OH | Solids | 47 | 174,000 |
| Pennsylvania | Chester, PA | Liquids | 44 | 220,000 |
| | Harrisburg, PA | Liquids | 51 | 125,000 |
| Texas | Gainesville, TX | Liquids | 39 | 17,300 |
| | Garland, TX | Solids | 48 | 200,000 |
| | Hollywood Road, Amarillo, TX | Liquids | 48 | 60,000 |
| | River Road, Amarillo, TX | Liquids | 43 | 140,000 |
| | South, Laredo, TX | Liquids | 42 | 120,000 |
| | Sunnyvale, TX | Solids | 48 | 186,000 |
| | Wichita Falls, TX | Solids | 49 | 90,000 |
| | Woodlands SJRA WWTF No. 1, TX | Liquids | 21 | 65,000 |
| | Woodlands SJRA WWTF No. 2, TX | Liquids | 21 | 70,000 |

| State | Plant | Sample Type | Sample Count | Population Served |
|---|---|---|---|---|
| | Woodlands SJRA WWTF No. 3, TX | Liquids | 21 | 15,000 |
| | Zacate Creek, Laredo, TX | Liquids | 41 | 140,000 |
| Utah | Central Salt Lake Valley, UT | Solids | 50 | 600,000 |
| | Provo, UT | Solids (Settling w Imhoof cones onsite) | 33 | 115,000 |
| Vermont | Essex Junction, VT | Solids | 16 | 30,000 |
| | Montpelier, VT | Solids | 15 | 10,100 |
| Virginia | Aquia, Stafford, VA | Solids | 17 | 100,000 |
| | Hillsville, VA | Solids | 49 | 3,000 |
| | Little Falls Run, Stafford, VA | Solids | 16 | 50,000 |
